# INDUCTION AND ORIENTATION PRACTICES AND PERCEIVED EFFECT ON HEALTH WORK FORCE PERFORMANCE AND SATISFACTION IN PUBLIC HOSPITALS OF ILUBABOUR ZONE, OROMIA REGIONAL STATE, SOUTH WEST ETHIOPIA, 2018: FACILITY BASED CROSS SECTIONAL STUDY DESIGN WAS EMPLOYED

**DOI:** 10.1101/2020.08.03.20165001

**Authors:** Sisay Siraj Ayana, Tesfamichael Alaro, Feyera Gebissa

## Abstract

**Background:** Successful orientation speeds up the adaptation process by helping new employees feel comfortable in the organization and by making them more productive on the job. In today’s world most organization are recognizing that this type of simple generic orientation is not enough, a more complex employee orientation or on boarding process is required.

**Objective:** To assess induction and orientation practices and perceived effect on health work force performance and satisfaction in public hospitals of Ilubabour zone, Oromia regional state, south west Ethiopia, 2018.

**Methods:** A Facility based cross sectional study was conducted by using both quantitative and qualitative methods. Quantitative data was collected by self-administered questionnaire and qualitative data was collected by in-depth interview with purposefully selected key informants using interviewer guide. The sample sizes for quantitative data were calculated by using single population proportion formula. Accordingly, a total of 403 samples were drawn from the source population by stratified sampling technique. Qualitative data of in-depth interview was transcribed, and thematically analyzed and triangulated with quantitative findings.

**Results:** Out of 388 only 135(34 %) respondents have attended induction and orientation training while assignment to different new responsibilities. The finding from key informants revealed that they take even for less than one day or an hour. Regarding the practices held during the training; only proper welcoming them to the organization and department was practiced, the rest practices like sharing the organization vision, involving senior leaders and post training evaluation was responded that they were exercised poorly within the organizations. Only 53.1% responded that induction and orientation training have perceived effect on employee Performance while 46.7% of respondents do not. About 55.7% respondents said that induction and orientation training have perceived effect on employees’ job satisfaction and the remaining responded to have no effect on employee’s satisfaction.

**Conclusion:** Generally, the induction and orientation practice was poor so as the supportive supervision and post training evaluation. More than half of the respondents perceived that it has effect on performance and satisfaction and the rest did not understand its specific effects.

## Introduction

Human Resource Management’s (HRM) effectiveness has been considered as a determinant of organizational performance. Which intern influenced by deliberate strategy in implementing and monitoring activities related to human resource management(1).among different HR practices; induction and orientation is the major ones that should be given for newly assigned employee to different position or works. Orientation is a process of integration of new employees into an organization. It helps new employees adapt to the work environment and their jobs. Orientation is a training opportunity to promote organizational effectiveness from the start of a person’s employment(2).

First impressions are to shape an individual’s image of the organization throughout their employment. New employees are already facing an anxiety inducing situation due to coming into a new environment and wondering if he or she will fit in. As such, it is important that the employer does not worsen the experience with a boring, confusing and overwhelming orientation process. It is important that new employees quickly feel like they belong and are a valued member of the organization(3).

Induction ensures a new employee is provided with information and assistance when commencing employment with an organization. Clearly outlining what the organization stands for and requires, reducing the risk of regulatory breaches and enabling employees to respond effectively to new responsibilities. Induction as the process of receiving employees when they begin work, introducing them to the organization and their colleagues, and informing them of the activities, customs and traditions of the organization(4). Induction has benefits for all involved in the process. Employees who settle quickly into their new job will become productive and efficient at an early stage and in turn will experience feelings of worth and satisfaction(5).

The occupation health and safety/OHS/ defines new employees as any person who is new to a position or place of employment, returning to a position or place of employment in which hazards have changed during the employee’s absence, under 25 years of age returning to a position or place of employment after an absence of more than six months and a person affected by a change in the hazards of a place of employment(7).

Many organizations consider recruiting to be more important than induction and thus the induction process is often neglected. However, it should be acknowledged that investing in recruiting will not pay off if the employee will not be committed through the induction process. Induction is often carried out during the work routines and many organizations assert that “you learn the best by doing”. It is also very common when hiring an already experienced employee the induction process is assumed to be less important.(8)

Employee orientation should set the tone for a long-lasting relationship between the employee and the organization. All too often, the practice is in such a hurry to put the new employee to work that key elements of the orientation are either ignored or delayed, creating a gap in the employee’s knowledge of the practice and contributing to a limited assimilation into the new culture.(9)

The induction and orientation programme instituted for newly assigned health work forces in the Ilubabour zone facilities Province may not be functioning effectively. As the zone is at remote area from the center newly assigned employees are very anxious when they took lottery by chance for deployment. Not only during the first joining time but they are urged to get transfer before they give service of one year.

The Oromia Regional Health Beraue has been insisting on induction training to the new employees. The region issued Secular guideline which directs all HWF/health workforces/employees to ensure that new employees are provided with opportunity to attend induction training(10).In most health sector no research conducted to assess the induction and orientation practices and its effects on newly assigned HWF in general and study area in particular. Therefore, this study aims to study the induction and orientation practices and its effects on HWF working in public hospitals Ilubabour zone (11).

## Methods

### Study area and period

An institution based cross-sectional study design was employed The study was conducted in two public hospitals namely Metu Karl referral and Darimu district hospital. Metu town is the capital city of Ilubabour zone which is located in South west Ethiopia, 600 KM away from Addis Ababa. According to woreda base health sector plan (WBHSP) population estimation report of FMOH Ethiopia, Ilubabour zone have estimation of total population by conversion factor it was around **933**,**325**during the time of the period 2010EC.This zone has public and private health facility currently providing different services for the community. There are 263 rural and 23 urban health posts,39 health centers, 104 private clinics11 rural drug venders and 30 drug stores are providing health care services. The study was conducted from August 13 to September 2, 2018.

### Study design

An institution based cross-sectional study design was employed in which both quantitative and qualitative research method was used.

### Source population

All health work forces within the Public Hospitals of Ilubabour zone were considered as source population.

Qualitative data: purposely selected four HR managers and two-line managers /Hospitals CEO/ from the selected hospitals and 2 HR managers and one-line manager from zonal health department. In depth interview was continued until saturation.

### Study population

All new health work forces recruited in 2010EFY, all HWF join the hospitals by transfer and HWF promoted within the selected health facilities and HWF assigned to new departments are comprises the study population.

### Sample Size Determination and Sampling Procedure

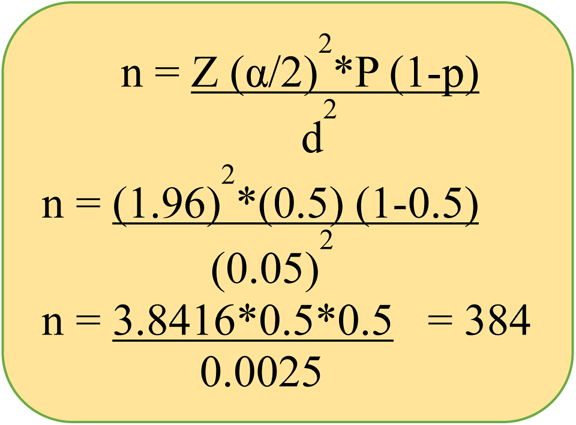

P =50%

CI=95 %(1.96). and 5 % degree of precision

Non response rate = 5%

n= 384

n=384+5% non-response rate

n=403

### Sampling procedure

All health work forces at MKH and DH hospital was stratified based on their profession (job) and **simple random sampling technique** was used to determine the sample size from each stratum. 40 new supportive staff and 42 health professionals was joined,7 supportive staff and 24 health care workers was joined by transfer and 3 health care professionals assigned by demotion totally health work forces were included purposely to the study and the rest individuals 136 supportive staff and 64 health care workers totally 100 was randomly selected from individuals promoted 2010 and 87 health care professionals were randomly selected from health care professionals changed their department in the same year from each stratum.

### Inclusion and Exclusion criteria

All health work forces present during the data collection time was included and health work forces that are recruited temporarily/daily laborer/ was excluded.

#### Data collection instruments

Quantitative data was collected by structured questioner and self-administered. The study instrument or questionnaire consists of three parts. Part one is about the demographic information including respondent, gender, job, educational level, experience, was included. Part two was about the induction and orientation practices and part three was on common perceived effect of induction and orientation program on employee performance and employee satisfaction. Furthermore, the qualitative data was collected by in-depth interview guide with purposefully selected key informants.

#### Variable for study

Induction and orientation training practices **Variables for descriptive analysis**

- Age - employee performance
- Gender - Employee satisfaction
- Job
- Educational level,
- Working department
- Benefit package,

#### Data Quality management

Data was collected by 4 trained data collectors and 2 supervisors who have an experience on datacollection and supervision and properly speak the local language and know the culture of the study population. Data collectors were trained by the principal investigator for two days about the objectives and purposes of the study and smooth ways of data collection. The questionnaires were designed from similar studies and translated to local language afan Oromo. To check the tool completeness and clarity pretest was done on 5%(17 individuals) of the total selected samples for study, and the pretest was done in Metu health center which is out of the study facilities. The collected data was entered to 3.1 version Epi data software for further data redundancy and quality control.

To maintain the qualitative data, the interview was done first after explaining the objective of the study and asking the key informants permission for recording their voice to make simple during transcription and to address well the messages missed on the time of note taking.

#### Data analysis

The quantitative data was cleaned, and finally exported to SPSS version 20 for analysis. Descriptive analysis (frequency, proportions, mean and standard deviations) were computed. Results were presented with tables, graphs and narrative presentations. Furthermore, qualitative data was transcribed, translated and analyzed using thematic analysis technique manually and finally triangulated with the quantitative findings.

### Ethical consideration

Ethical clearance was obtained from the institutional Review Board (IRB) of Jimma University, institute of Health Science. the Ilubabour zone health office based on the JU cooperation letter second cooperation letter was also obtained that directed to hospitals.

#### Operational definition

Satisfaction: It is a satisfaction level of individuals measured with 5 level of Likert scale in line to induction and orientation training. the result was discussed if the mean score is above the average they are satisfied and if it’s under the mean unsatisfied.

## Result

### Socio-demographic characteristics

In this study out of the total 403 sampled health workers, 388 health workers were studied with overall response rate of 96%. Out of the total studied health workers 283(70.2) and 120(29.8) were from Metu referral Karl hospital and Darimu district hospital respectively. In this study more than half of respondents 209(53.9%) were males. The dominance number of respondent age group was 20-29years 238(61.3%). Most respondents were from administration 45.6 %(177) followed by inpatient departments 28.4 %(110) while the third was out patient department 26 %(101). The distribution of departments might be attributed to the interplay of several factors such as the management perception of the need for induction orientation, internal directive, job design, job rotation. Other factors in department manner are specially IPD and OPD had different reforms and new initiatives are implemented and closely need induction and orientation for new comer employees respectively. From the listed benefit packages for employee’s car service were the dominant benefit package 54.6 %(212). The next dominant benefit package was having own office and benefit from private wing service are 22.4 %(87) and 12.4(48) respectively. The rest benefit packages are housing and telephone service benefit package which accounts 7.7 %(30) and 2.8(11) respondents respectively.

### Induction and orientation training attending status of employees

According to respondents answered over question asked on having induction training only 34.7%(135) respondents were attended induction and orientation training within their current organization and majority of the respondents 65.3 %(253) were not attended. from though having induction and orientation training 40%(54) of them attended orientation during department changes,31.9%(43), during promotion,17%(23), during first appointment,9.6%(13), during transfer from different organization to hospitals and 1.5%(2) attend induction and orientation training during demotion. As the respondents responded on duration of the induction and orientation training 75.4 %(102) were attended only for a day and 17.9 %(24), 5.2 %(7) and 1.5%(2) respondents responded that two, three and a week duration respectively.

### Induction and orientation training practices

#### Practice 1. Engagement

practice concerning engagement of new employee with others during Orientation were only 39.2% of total respondent said the orientation create interesting and its beyond lecture to stimulate knowledge transfer, the rest 60.8% were responded there in no creating interested during induction and orientation, when the respondents were asked for whether during the training role play and team game were part of the training they were respond that, 77.3%(300) of them said there were no role play and team game during the training and 22.7(88) of them responded that there were such practice during the training. practice concerning engagement of new employee with others during Orientation were only 39.2% of total respondent said the orientation create interesting and its beyond lecture to stimulate knowledge transfer, the rest 60.8% were responded there in no creating interested during induction and orientation, when the respondents were asked for whether during the training role play and team game were part of the training they were respond that, 77.3%(300) of them said there were no role play and team game during the training and 22.7(88) of them responded that there were such practice during the training.

#### practice 2. Welcoming

Respondents were asked whether they were provided with welcome celebration activity like an organization -wide welcome party or departmental level welcoming their response were indicated that,52.8%(205) of them are welcomed by the organization and department level and the rest 47.2%(183) of them were not welcomed by the organization as well as by department. As respondents were asked whether they were Provided with detailed tours of the facility the table 3 shows that, 26.5%(103) were provided with tour of the facility or the department the rest 73.5%(285) were responded that not provided with tour of facility. During welcoming practice respondents were asked whether they were joined with provision and arrangement of many productivity tools and other assets for them to be available on day one as possible, the result above shows that 63.4%(246) of them responded it was providing for then and the rest 36.6%(142) of them responded that it was not provided with arranged tools and assets on the first day of joining and for question they were asked on whether the respondents were got orientation or training on the tools and assets, 53.1%(206) of respondents responded they were taking orientation and training and the test 49.9% of respondent answered they don’t have orientation or training over the tools and assets.

**Table 1:** Socio-demographic characteristics of health work forces in in Metu Referral Karl and Darimu Hospital, 2018(n=388)

|  | Frequency | Percent |
| --- | --- | --- |
| **1.Sex of respondents** |  |  |
| male | 209 | 53.9 |
| female | 179 | 46.1 |
| Total | 388 | 100.0 |
| **2.Age in year** |  |  |
| <20 | 4 | 1.0 |
| 20-29 | 238 | 61.3 |
| 30 to 39 | 100 | 25.8 |
| 40-49 | 34 | 8.8 |
| >50 | 12 | 3.1 |
| Total | 388 | 100.0 |
| **3.Working department** |  |  |
| outpatient department | 101 | 26.0 |
| inpatient department | 110 | 28.4 |
| Administration staffs | 177 | 45.6 |
| Total | 388 | 100.0 |
| **4.benefit package** |  |  |
| Housing subsidy | 30 | 7.7 |
| Car allowance /service | 212 | 54.6 |
| Own office | 87 | 22.4 |
| cellphone | 11 | 2.8 |
| Private wing | 48 | 12.4 |
| Total | 388 | 100.0 |

**Table 2.** Induction and orientation training attending status of health workforces in MKH and DH, 2018

| 1.Induction ad orientation training attending status |  |  |
| --- | --- | --- |
|  | Frequency | Percent |
| yes | 135 | 34.7 |
| no | 253 | 65.3 |
| Total | 388 | 100 |
| 2.the time when they have induction and orientation training |  |  |
|  | Frequency | Percent |
| First Appointment | 23 | 5.9 |
| During change of Department | 54 | 13.9 |
| Promotion | 43 | 11.1 |
| Demotion | 2 | 0.5 |
| transfer | 13 | 3.4 |
| Total | 135 | 34.8 |
| Total | 135 | 100 |
| 3.Duration of induction and orientation training |  |  |
|  | Frequency | Percent |
| One day | 102 | 26 |
| Two days | 24 | 6.2 |
| Three days | 7 | 1.8 |
| Week | 2 | 0.5 |
| Total | 135 | 34.5 |
| Total | 135 | 100 |

**Table 3:** Responses of Health workforces on engagement practices during induction and orientation practices in Metu Referral Karl hospital and Darimu hospital,2018

|  | Frequency | Percent |
| --- | --- | --- |
| 1. create interesting beyond lecture to stimulate knowledge transfer |  |  |
|  | Frequency | Percent |
| yes | 152 | 39.2 |
| no | 236 | 60.8 |
| Total | 388 | 100.0 |
| 2. role play and team game during induction and orientation training |  |  |
|  | Frequency | Percent |
| yes | 88 | 22.7 |
| no | 300 | 77.3 |
| Total | 388 | 100.0 |

**Table 4:** Responses of Health workforces on welcoming practices during induction and orientation training in Metu Referral Karl hospital and Darimu hospital 2018

|  | Frequency | Percent |
| --- | --- | --- |
| **1. celebration activity like organization -wide or departmental welcome party** |  |  |
|  | Frequency | Percent |
| yes | 205 | 52.8 |
| no | 183 | 47.2 |
| Total | 388 | 100.0 |

| **2. detailed tours of the facility** |  |  |
| --- | --- | --- |
|  | Frequency | Percent |
| yes | 103 | 26.5 |
| no | 285 | 73.5 |
| Total | 388 | 100.0 |
| **3.productivity tools and other assets to be available on day one as possible** |  |  |
|  | Frequency | Percent |
| yes | 246 | 63.4 |
| no | 142 | 36.6 |
| Total | 388 | 100.0 |
| **4. training on these tools and assets** |  |  |
|  | Frequency | Percent |
| yes | 206 | 53.1 |
| no | 182 | 46.9 |
| Total | 388 | 100.0 |

**Table 5.** Responses of Health workforces on involvement of senior leaders during induction and orientation training in Metu Referral Karl hospital and Darimu hospital.2018

|  | Frequency | Percent |
| --- | --- | --- |
| **1.Involvement of senior leadership in induction and orientation training programs** |  |  |
|  | Frequency | Percent |
| yes | 99 | 25.5 |
| no | 289 | 74.5 |
| Total | 388 | 100.0 |
| 2. executives to present a portion of the program |  |  |
|  | Frequency | Percent |
| yes | 70 | 18.0 |
| no | 318 | 82.0 |
| Total | 388 | 100.0 |

As subjects generally replied, induction and orientation program was one program from HR practices but as other HR practices it doesn’t have attention like others programs due to different reasons like program overlap, budget allocation for the program, time of new employee joining to the sector and awareness on the importance of the program are problems as the key informants replied respectively.

A 48 years old man from one public hospital said that, “*since I have a teaching experience for a long time induction and orientation is common in education sector. but when am joining this hospital am only introduced to my coworkers and received material which are regulated under the HR department I don’t received any induction and orientation training during am assigned in this hospital. since the organization was wide and complex by discussing with CEO and matron in our organization when new comers joined our hospital introducing with department heads and coworkers was done and to some extent orientation on the rule and regulations of the hospital a few orientations how they keep in the hospital service provision system we were give orientation but it is not well organized as that of education sector*.

### Practice 3 involve senior leaders

As indicated on Table 6 below, majority 74.2% (289) of respondents answers that senior leaders were not Involved in induction and orientation program and the rest of respondents 25.5%(99) were answered that senior leaders were involved during induction and orientation training programs. As the respondents were responding that during induction and orientation program leaders of the hospitals had not participated by presenting portion of presentation during new comers joined the hospitals, the dominants 318(82%) of respondents answer that no senior leaders were involved in induction and orientation training with involvement on portion of presentation and the rest 18%(70) respondents answer that senior leaders present a portion of induction and orientation presentation for new comers.

**Table 6.** Responses of Health workforces on shared vision, mission and goal of the organization during induction and orientation training in Metu Referral Karl hospital and Darimu hospital,2018

|  |  |  |
| --- | --- | --- |
|  | Frequency | Percent |
| 1. introducing them to health care organization history, mission, and values during the induction |  |  |
|  | Frequency | Percent |
| yes | 125 | 32.2 |
| no | 263 | 67.8 |
| Total | 388 | 100.0 |
| 2. Helping new employees understand the core programs of the institution through skilled training |  |  |
|  | Frequency | Percent |
| yes | 139 | 35.8 |
| no | 249 | 64.2 |
| Total | 388 | 100.0 |

#### practice 4 shared vision, mission and goal of the organization

As indicated on table below, majority of the respondents 67.8% (263) were responded that during induction and orientation training program introducing them to health care organization vision history, mission, and values were not created a pride to new comers and the rest respondents 32.2%(125) answered that they were introduced with the organization, vision, mission, history and values during the induction and orientation training program. As the respondents responded over orientation on core program of the institution, majority of the respondent 63.9%(249) were respond that they were not oriented on core programs of the institution through skill training and an overview of the lines of work flow and the rest 35.8% (139) were responded that they were oriented on core programs of the institution through skilled training and overview of the lines of work flow.

A 28 years old man who was from one hospital said that, *“as think am not sure whether my organization mission and vision was 100% well interpreted in all level of my organization employees. I mention above that my organization have orientation program but not well organized induction program was given for newly assigned employees this may be the failures but we have clear mission and vision indicating tapela /brochure outside at the door of my office*.”

A 30 years old man from one public hospital said that, *“I think am not confident to say the organization mission and vision is well interpreted, but during welcoming new employees with presentation of right and responsibility of employees we highlight the mission and vision of the organization, to give in detail there was a problem of budget*.*”*

### practice 5 post training evaluation

#### 5.2.5. practice 5 post training evaluation

Since induction and orientation program is a continuous program it need post training evaluation how the program helps the new comers throughout their career. As Pie chart above shows majority of the respondents 53.1%(206) respond that there were no a structured follow-up system involving HR/training, hiring managers, and a facilitated mentoring process, the rest respondents 46.9% (182) were respond that there were a follow-up system involving HR/training, hiring managers, and a facilitated mentoring process.

**Figure:**
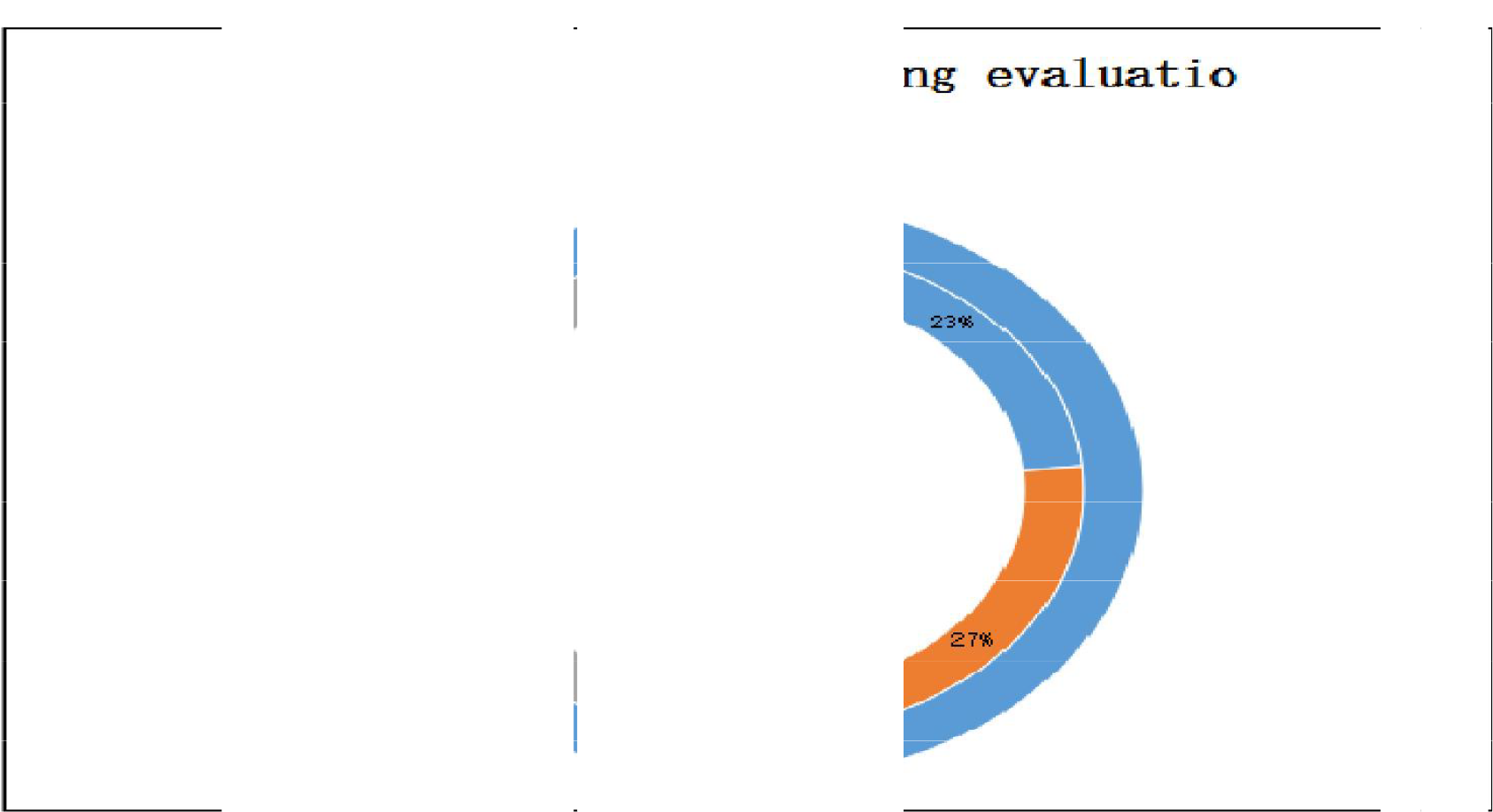
Post training evaluation,2018

### Perceived Effect of induction on employee performance

Out of the aggregated responses provided by respondents to the seven statements on the perceived effect of induction on performances, equal shares of 29% went to strongly agree (SA) and 39% agree. Neutral had 22%, Disagree (DA) 6%, and Strongly disagree (DSA) 4%. The statements “I am highly attracted to my work” (6.2%) as against “My work and its related activities are more important than others” (1.3%) turned out to be respective dominant and the least frequent that were rated ‘SDA’. In respect of ‘DA’ the first position went to “I am highly attracted to my work” (14. 4%) were dominantly rated and “I always finish assigned jobs within time” (2.6%) was least frequent rated. The last position went jointly to “I always feel bothered for job failures” and “I perceive the importance of being identified with my work and evaluating others worth on that basis” (8.2% each). The statement “I am highly attracted to my work” (36.1%) was accorded the highest neutral rating. On the contrary, “I always come to work on time” was accorded the lowest. The leading, ‘Agree’ statement was “I always come to work on time” (45.9%) while the least important was “I am highly attracted to my work”. Lastly, the leading ‘SA’ statement was “I always feel bothered for job failure” (37.9%) whilst the least prominent “I am highly attracted to my work” (16.5%).

A 28 years old man from zonal health department said that, “*yes it has an effect on performance this is because of if somebody have a well-organized orientation in any discipline it motivation will improved then if he is motivated and understand the way how he performs tasks the individual as well as the organization performance became improved, so it has an effect over performance*.*”*

In organization where there is poor practice of induction and orientation for new comer employees rather than early engaging to the organization overall activities sometimes it may have negative consequence over the new comer as well as over the organization.

48 years old man from one public hospital said that, “*For example, last year when we recruited cleaners to this hospital due to not provided with infection prevention training there was a problem of exposed to some injury to one cleaner. But currently we are trying to orient them by trained professionals for a few hours on the first day of joining the hospital”*.

### Perceived Effect of induction and orientation on employee job satisfaction

Out of the aggregate of 377 responses provided by respondents to the ten statements on their job satisfaction, “agree” topped with 31.2%. “neutral” followed with 27% satisfied, while “Dissatisfied” came third with 19.22%. The fourth position went to ‘greatly dissatisfied’ (12. 26%).The least frequent assessment were 26 ‘greatly satisfied’10.41%.

In the case of “greatly dissatisfied”, the top position went jointly to salary, which is followed by the lowest rated variable Your overall satisfaction with your organization with 3.4%. In terms of the “Dissatisfied” Benefits, allowances and bonuses 40.5% was dominant. The least important variable was overall satisfaction with an organization 6.4% followed by relationship with your peers (7.2%).

In the case of “neutral”, the top position wants jointly to job security (38.4%) while the least position went to salary (17. 5%).The first and second positions on the “satisfied” rating went to Your overall satisfaction with your organization (43%)followed by Recognition received from your supervisor and relationship with peers (36%).The least position went jointly to salary 7.7% and Benefits, allowances and bonuses 11.1% respectively.

### Challenges in induction and orientation practices

The challenges in induction and orientation practice for employees in the study institutions include the planning of the curricular, budget allocation problem, training, monitoring and evaluation and well developed guideline are challenges in induction and orientation program.

A. Planning: Induction and orientation program is one program from human resource practices that need special attention in succeeding organizational goal. To implement induction and orientation program in an organization the initial task is to be planned as a program with others human resource management program is very important. But currently, within the study conducted hospitals paper wise with other HR programs and activities, with clear time frame and clear implementation strategy there is no plan developed on induction and orientation program.
B. Budget allocation: Since induction and orientation program have an importance in early engagement of employees to an organizational culture before joining an organization as well as before being assigned to new work area having an induction and orientation is important. To give well organized training budget allocation and well utilization have an effect on equipping the new employee for the new task. A 28 years old man from at zonal health department said that, *“starting from higher leaders the program doesn’t get enough attention as that of other program and the other is as this program is very important but there is no budget allocated in special case for induction and orientation program to give more than one day*.” A 48 years old man at one hospital said that, “*As I think there are challenges like budget constraint, work over lap/being busy for unplanned task/ are the major challenges in practicing induction and orientation program*.*”*
C. Training: To give successful induction and orientation training enough preparation with suitable training environment is important with its good consistency of presentation. Both hospitals during they welcome new employee rather than giving well reached orientation they focus mainly on rules and regulation like work time explanation and introducing to their department colleague. A 30 years old man who from one public hospital said that, “*Regarding challenges faced with conduction of induction and orientation training budget for the program is first challenge for me, the other is individuals who get training they didn’t give for the others at work area. The other is well organized presentation was not prepared due to the is no well-developed and updated guideline over the program*.*”*
D. Monitoring and Evaluation. Induction and orientation is a very important program in any organization but specially in health sector was it doesn’t get enough attenuation like other HR programs.so as key informants said during interview the program doesn’t get emphasis starting from planning to monitoring and evaluation. During supervision was given there is nothing indicator related to induction and orientation included in supervision checklist.

A 30 years old man from one public hospital said that,” *I don’t have any supervision took on induction and orientation program as well as on other HRHM practices, you know our hospital is at remote area. But the concerned body should be give emphasis for the program and for other services*.*”*

A 28 years old man from one public hospital said that,” I have supervisory support from region but it is not regular and specially in respect to the case you ask me I didn’t get supervisor *support from anybody previously but as much as possible we are trying to give orientation for new comers*.*”*

Finally, findings from in-depth interview revealed that induction and orientation program was practiced with different constraints that gone to different stake holders. The first problem in line to the program was it doesn’t have integrated plan with others human resource management practices. As key informants discussed if the program were not having attention clearly starting from the higher managers to the grass root level the implementation became un effective and it puts negative effect over the organizational goal. The next problem raised by the interviewee was standardized guideline for the program were critical. The other problem was problem of budget allocation for the program to give detail knowledge and skill for newcomers during induction ad orientation training provided beyond an hours or a day.

## Discussion

As discussed before in different part of the research body specially in the health sector enough research’s on induction and orientation program were not conducted specially there were a very scarce of reference to discuss detail on the program as well as with the findings from this research.

Induction and orientation program equip the new entrants with basic knowledge and skill is ineffective but less efficient. From overall newly assigned health work forces only 34.7% of them assigned with having induction and orientation. when its analyzed with the guideline on Induction and orientation training should be provided for all newly assigned HWF it was practiced poorly.

An induction program is part of an organizations knowledge management process and is intended to enable the new comers to become a useful and integrated member of the team. An organization should acquaint new employees’ technical and theoretical knowledge necessary to do their job efficiently or how their role fits in with the rest of the organization. It is essential that employers should educate employees regarding their role, key result areas and organizations expectations in advance to curb attrition at the later stage during a skills dialogue session. It will help employees to understand why they are hired and how they contribute to the success of the organization goals. The findings from these study on engagement of new employees reveal that 39.2% of total respondent said the orientation create interesting and its beyond lecture to stimulate knowledge transfer, the rest 60.8% were responded there in no creating interested lecture during induction and orientation. The overall goal of induction is to help new employees learn about the organization as soon as possible, so that they can begin contributing.

Induction and orientation program gives new entrants an overall insight regarding the organization Mission, vision, values as well as about the institution-wide culture. It builds culture and vision. The induction and orientation training program has a vast contribution to comprehend new comers the vision mission values and the overall culture of the organization. It’s known that an organization cannot fully achieve its reasons for existence without a proper definition of its vision and mission statements, hence vision and mission statements can be said to be a catalyst or driving force in any organization activities as Understanding the mission statement is very significant to an employee’s continued success and happiness of the organization. But, findings from study reveal that 67.8% (263) were responded that during induction and orientation training program introducing them to health care organization vision history, mission, and values were not created a pride to new comers and the rest respondents 32.2%(125) answered that they were introduced with the organization, vision, mission, history and values during the induction and orientation training program and the finding from two key informants revealed that there were poor interpretation of vision and mission practiced during inducing new employees to the organizations P1 said that,*“as think am not sure whether my organization mission and vision was 100% well interpreted in all level of my organization employees. I mention above that my organization have orientation program but not well organized induction program was given for newly assigned employees this may be the failures but we have clear mission and vision indicating tapela /brochure outside at the door of my office*.” and P2 said that, *“I think am not confident to say the organization mission and vision is well interpreted, but during welcoming new employees with presentation of right and responsibility of employees we highlight the mission and vision of the organization, to give in detail there was a problem of budget*.*”*

The findings from induction and orientation practices indicated that only during welcoming introducing the new comers to the organization wide or with departmental level employees and provision and arrangement of many productivity tools and other assets for them to be available on day one was the two activities well practiced during welcoming program. The rest four induction and orientation practices employee engagement, senior leader’s involvement, sharing the organization mission, vision of an organization and post training evaluation was practices poorly within the studied organization. These was also well discussed by the key informants during in depth interview were conducted with them.

Findings from effect of induction training on employee performance reveal that,53.1% of respondents was perceived that the induction and orientation training have effect on their performance and the rest respondents were perceived that they don’t know whether it have or not an effect on their performance and it indicated that there was no have well preparation and continuous evaluation. The other finding was the effect of induction and orientation training on employee’s job satisfaction the findings show that, **55.7%** of respondents was perceived that the effect of induction and orientation training have effect on their job satisfaction and the rest respondents were perceived that they don’t know whether it’s have effect or not on their satisfactions as that of effect on employee’s performances above.

Both findings were revealed that the induction and orientation training was practiced poorly within the studied organizations.

As that of other HR practices induction and orientation program doesn’t got enough attention like employee selection, recruitment, employee development, performance appraisal etc. when new comers join an organization and new position if they don’t have enough induction orientation training they became confused and anxious then it leads them to have poor performance and unsatisfied with their current work. The finding from qualitative result reveal that if well-organized induction ad orientation training was provided for new employees they assure its have positive effect in employees performance improvement as well as employees to be satisfied with their job respectively. from the key informants responses on effect of induction and orientation on employees performance and employee job satisfaction P1 said that, *“sure! it has an effect on employee performance as well as on over all organizational performance. Any employee if he/she has well organized induction and orientation training before his attachment for new tasks he/she will engage to the new task within a short time, this a short time engagement will lead the employee to have a good performance that supports the overall success of the organization goal respectively*.*”* and P2 said that, “*yes it has an effect on performance this is because of if somebody have a well-organized orientation in any discipline it’s motivation will be improved then if he is motivated and understand the way how he performs tasks the individual as well as the organization performance became improved, so it has an effect on employee performance as well as on their job satisfaction*. “

Finally, findings from the in depth interview was also strengthen the findings from quantitative result of induction and orientation practices and effect of the training on employee’s performance and employee job satisfaction as well.

The limitation of the study was Similar to other studies; this study has limitations of self-reported data bias. As the study design is cross-sectional it is difficult to determine exact cause and effect relationship among variables. Another limitation of this study was limited literatures to discuss in Africa and Ethiopia context.

## Conclusion

In general the induction and orientation practices assessed in this study was exercised poorly with poor supportive supervision and post training evaluation. Organizations were not sufficiently exert their effort to address the induction and orientation practices like employee engagement, well organized welcoming, senior leader involvement in the program, history, mission and vision of the organizations and evaluation post the training through a well-developed plan and orientation program for their new comers and current employees. As well discussed in discussion chapter majority of the employees perceived that the program has effect of their performance and satisfaction and the rest were not understanding whether induction and orientation training provided in their organization have effect on their performance as well as on their job satisfaction. These result was also supported by the qualitative findings from key informants responding that currently as that of other sector like education it doesn’t got enough focus but if it will get organized attention throughout the health sector structure it’s have a positive effect on performance improvement and employee job satisfaction. All the objectives set were achieved and with regard to the main objective of the study it can be concluded that the following problems, such as lack of well-developed guideline, lack of budget allocation for the program, unsuitable working environment inadequate logistics/materials etc. were indeed the key factors affecting effective employee induction and orientation in the studied organization.

## Data Availability

Data was collected by 4 trained data collectors and 2 supervisors who have an experience on data collection and supervision and properly speak the local language and know the culture of the study population. Data collectors were trained by the principal investigator for two days about the objectives and purposes of the study and smooth ways of data collection. The questionnaires were designed from similar studies and translated to local language afan Oromo. To check the tool completeness and clarity pretest was done on 5%(17 individuals) of the total selected samples for study, and the pretest was done in Metu health center which is out of the study facilities. The collected data was entered to 3.1 version Epi data software for further data redundancy and quality control.
To maintain the qualitative data, the interview was done first after explaining the objective of the study and asking the key informants permission for recording their voice to make simple during transcription and to address well the messages missed on the time of note taking.

## Acknowledgment

First of all, I gratefully thank Jimma University, institute of health, faculty of public health department of health economics, management and policy for giving me this incredible chance to prepare this research proposal.

Secondly, my special thanks also goes to my advisors, Mr. Tesfamichael Alaro (BSC, in PH, MHPE, MPH, assist proff.) and Mr. Feyera Gebissa (B. Pharm, MHA) for their unreserved help starting from title selection until maturation of this work.

Finally, my sincere thanks goes to Mr. Kebebe Bidira by sharing his experience on research preparation, my brother Michael Siraj, my family and all those, who in one way or another have contributed to the success of this research preparation.

## Competing interest

I declare that there is no both financial and non-financial competing interest.

## Notes

### Competing Interest Statement

The authors have declared no competing interest.

### Funding Statement

The funding organization was Jimma university and the funder have no any role in the design, data collection, analysis and publication of this paper.

